# Recurrence patterns in a large contemporary cohort of patients with non-muscle invasive bladder cancer

**DOI:** 10.1101/2024.12.19.24319161

**Authors:** Jasper P. Hof, Lambertus A. Kiemeney, Katja K.H. Aben, Antoine G. van der Heijden, Alina Vrieling, Sita H. Vermeulen

**Affiliations:** Radboud university medical center, IQ Health science department, Nijmegen, The Netherlands; Radboud university medical center, Department of Urology, Nijmegen, The Netherlands; Netherlands Comprehensive Cancer Organization, Department of Research and Development, Utrecht, The Netherlands

**Author notes:** These authors share senior authorship (Alina Vrieling, Sita H. Vermeulen).

**Keywords:** bladder cancer, recurrence, progression, recurrent event analysis, risk stratification

## Abstract

**Background and Objective:** Patients with non-muscle invasive bladder cancer (NMIBC) frequently experience recurrences, yet, the timing and characteristics of subsequent recurrences are understudied. We aim to describe subsequent recurrences in a large, contemporary, population-based cohort.

**Methods:** We included 1,915 patients from the UroLife study and the Nijmegen Bladder Cancer Study, diagnosed with primary NMIBC between 2011 and 2021. The conditional 1-, 3- and 5-year risks of first to fourth recurrence were calculated using Kaplan Meier risks, stratified by clinicopathological factors. Patterns of subsequent tumours were described and visualised.

**Key Findings and Limitations:** We observed 671 first recurrences and 400 subsequent recurrences. The 3-year conditional recurrence risks for first, second and third recurrence were 31%, 45% and 54%, respectively, and were similar for NMIBC risk groups. Recurrence after a low-, intermediate- or high risk tumour (either primary or recurrent) was again of low-, intermediate- or high-risk in 74%, 62% and 44% of cases, respectively. Ten patients with low/intermediate risk NMIBC and 89 patients with high risk NMIBC progressed to muscle-invasive or metastatic bladder cancer. Seven out of these ten (70%) and 13 out of these 89 (15%) patients had a high risk recurrence before progression. This study was limited to a patient cohort from The Netherlands.

**Conclusions and Clinical Implications:** Our study provides novel and reliable estimates of recurrence rates and patterns in NMIBC from a large, contemporary, population-based cohort. Recurring NMIBC tumours often exhibit similar clinicopathological characteristics, and patients with primary low- or intermediate-risk NMIBC who progress to MIBC/mBC often have a high-risk recurrence before progression. These results can inform research into NMIBC recurrences and surveillance schedules.

**Patient Summary:** Our research focused on the timing and characteristics of recurring tumours among patients with non-muscle invasive bladder cancer. We showed that a recurring tumour is often of a similar type as the previous tumour. We also found that for patients with initial low- or intermediate risk tumours, a progression to muscle-invasive bladder cancer is often preceded by a high risk recurrence.

**What does the study add?:** Existing literature extensively addresses the risk of first recurrence as clinical outcome, leaving the risk, timing and characteristics of subsequent recurrences relatively understudied. Our study addresses this gap by investigating patterns of subsequent recurrences in a large contemporary population-based cohort.

We found that recurrence risk increases with number of prior recurrences, and subsequent tumours often manifest similar clinical characteristics within patients. Moreover, we discovered that over half of patients with primary low- or intermediate risk NMIBC who progress to muscle-invasive or metastatic disease experience a high risk recurrence before progression.

## Introduction

Approximately 75% of patients with bladder cancer are diagnosed with non-muscle invasive bladder cancer (NMIBC), which is generally associated with a favourable survival (1–3). Yet, patients with NMIBC face a high risk of recurrence and a substantial risk of progression to muscle-invasive bladder cancer (MIBC) or metastatic disease (mBC) (1), necessitating an intensive follow-up schedule with frequent invasive cystoscopies (2, 3).

Extensive research has been conducted into the clinical and pathological characteristics that predict NMIBC recurrence and progression (4–11). Resulting risk classification models that classify NMIBC tumours into low-, intermediate-, high-, or very high risk are used in clinical practice (3, 12). However, most studies primarily focused on the first recurrence as clinical outcome, while the risk and timing of subsequent recurrences remain relatively understudied. Yet, subsequent recurrences have been reported to comprise 47% up to 62% of all observed recurrences (11, 13–16).

Although prior recurrence is a predictor in NMIBC risk classifications (5, 14), data on risks of subsequent NMIBC recurrences are limited (11, 14, 15, 17, 18), and no study has explored the patterns of tumour characteristics of these recurrences, including risk group classification. Previous studies suffer from limitations such as the use of data that can now be considered outdated (14, 16), relatively small sample sizes (15, 17), or restriction to an intermediate risk group (17). A more comprehensive understanding of contemporary patterns of NMIBC recurrences may facilitate research into mechanisms of recurrence and progression, improved risk classification, and novel treatment and follow-up schedules.

In this study, we evaluated the timing and characteristics of recurrences of patients diagnosed with NMIBC. All analyses were performed for the total cohort and stratified by risk group of the primary tumour, which was based on a modified version of the EAU risk classification. We used two large population-based Dutch bladder cancer cohorts: the UroLife study and the Nijmegen Bladder Cancer Study (NBCS), in which recurrences were systematically registered, providing a robust foundation for our investigation.

## Materials and Methods

### Study population

Patients diagnosed with NMIBC were selected from two large Dutch bladder cancer cohorts. Both cohorts identified eligible patients through the Netherlands Cancer Registry (NCR), which records data from all cancers diagnosed in The Netherlands since 1989. One cohort is the UroLife study (19) (N = 1,515), which is a prospective multicenter cohort study conducted in 22 hospitals in the east, south and central parts of The Netherlands. Eligible patients were between 18 and 80 years old, underwent a TURBT, and were newly diagnosed with histologically confirmed primary NMIBC of stages Ta (May 2014-April 2017), T1 (May 2014-April 2021), or Tis (May 2014-April 2017). Patients were invited for participation approximately 4 weeks after diagnosis. Response rate was 52% and participants were comparable to patients declining participation with respect to age, sex, tumour stage and tumour grade. Ethical approval was provided by the Committee for Human Research region Arnhem-Nijmegen (CMO 2013-494).

The other cohort is the Nijmegen Bladder Cancer Study (20) (NBCS). During the period 1995-2015, patients diagnosed with bladder cancer in one of seven hospitals in the eastern part of The Netherlands were enrolled in the study. Patients were considered eligible if diagnosed at or before the age of 75, alive at time of invitation for the study and able to read/understand the Dutch language. Invitation was performed in six separate phases or batches. For the current study, we only used the three most recent NBCS batches with patients who were diagnosed between 2011-2015 and invited for participation in 2017 and 2019 (N = 472). The NBCS was approved by the Committee for Human Research region Arnhem-Nijmegen (CMO 2005-315).

Patients who participated both in UroLife and the NBCS were identified (N = 36) via linkage to the NCR database and the duplicate record with the shortest follow-up time was omitted. Patients with upper tract urothelial carcinoma before (N = 10) or within 3 months of primary diagnosis (N = 26) were excluded. For both UroLife and NBCS, relevant data for this study on patient (age, sex) and tumour characteristics (tumour stage, tumour grade, CIS, focality), and outcomes were obtained by a medical file review by NCR data managers.

Treatment and follow-up of patients with NMIBC in The Netherlands is according to the Dutch Urology Association (NVU) guidelines, which are based on the EAU guidelines. However, urologists may deviate from guidelines in their clinical practice.

### Definition of recurrence and progression

Recurrence is defined as either a (i) histologically confirmed bladder tumour based on a trans-urethral resection of bladder tumour (TURBT), (ii) report of fulguration, (iii) histologically confirmed bladder tumour found after cystectomy, or (iv) progression to MIBC or mBC after an initial NMIBC diagnosis. Fulguration only counted as recurrence if it occurred at least six months after previous recurrence, and was coded as TaG1 recurrence.

Notably, a recurrence can only occur after the patient achieved a ‘tumour-free status’. The definition for tumour-free status depends on the stage of the previous bladder tumour:

- For stage Ta or T1 tumours, an individual was considered tumour-free after undergoing radical TURBT or re-TURBT, or, in case of nonradical (re-)TURBT, after tumour-negative cystoscopy;
- For stage Tis or concomitant carcinoma in situ (CIS), an individual was considered tumour-free after tumour-negative cystoscopy, and, if available, negative cytology. In cases where a biopsy was performed, a negative result was required.

Notably, the requirement for negative cystoscopy and, if available, negative cytology and biopsies ensured that for (concomitant) CIS, only those patients who had a complete response after treatment were at risk of recurrences. As a result, 42 patients with primary (concomitant) CIS who were never at risk of recurrence were excluded from analysis of recurrence. However, they remained at risk of progression and were hence included in the progression analysis. Progression to MIBC or mBC was defined as (i) the occurrence of a bladder tumour of stage T2 or higher, confirmed by pathology or imaging, (ii) positive lymph node status, or (iii) distant metastasis.

Patients were no longer at risk of recurrences after either (i) the last contact between patient and urologist; (ii) radical cystectomy; (iii) progression to MIBC or mBC; or (iv) diagnosis of another type of cancer with metastasis, whichever came first. Patients were no longer at risk of progression to MIBC/mBC after either (i) the last contact between patient and urologist; or (ii) diagnosis of another type of cancer with metastasis, whichever came first. Recurrence-free survival was defined as the time between the date of achieving the tumour-free status and date of subsequent recurrence or end of at-risk period, whichever came first.

### Definition of risk groups

Bladder tumours were categorized based on the EAU risk group classification (2021) by Sylvester *et al.*, which is based on stage, grade, (concomitant) CIS, prior recurrence, focality, age, and tumour size (5). We however adopted a slightly modified version in which prior NMIBC recurrence was not included as risk factor, to ensure comparable classification of primary and recurrent bladder tumours (see **Supplemental Table 1**). Also, tumour diameter was omitted, since this was not systematically registered in the medical records by Dutch urologists and therefore mostly missing. Consequently, a small percentage of tumours was assigned to a lower risk group compared to the EAU risk group classification.

### Data analysis

We assessed the 1-year, 3-year, and 5-year risk of first to fourth recurrence, conditional on prior recurrence, based on Kaplan-Meier (KM) risk. To investigate whether recurrence patterns differ by recency of diagnosis, we also performed these analyses separately for patients with a primary NMIBC diagnosis before and after 2016 (median year of diagnosis). We also stratified KM analyses by treatment, as NMIBC prognosis is affected by treatment. In addition, we stratified the KM risks of first to fourth recurrence by sex to explore possible sex-specific recurrence patterns, as women have been reported to have poorer NMIBC prognosis (21).

Next, we described the transitions between risk groups, grade, stage, and tumour location across subsequent recurrences within patients. These were also visualised using alluvial diagrams. All KM risks and transitions between risk groups across subsequent recurrences were evaluated for the total cohort and stratified by risk group of the primary tumour. Finally, we described the recurrence patterns of the patients who progressed to MIBC or mBC using swimmer plots.

All analyses were performed in R (version 4.1.3). KM plots, alluvial diagrams and swimmer plots were visualised using the ggplot2 package (version 3.3.6).

## Results

### Cohort characteristics

Patient and tumour characteristics at time of primary diagnosis and at time of first to fourth recurrence are described in detail in **Table 1**. A total of 1,915 patients with a primary diagnosis of NMIBC were included, and 1,071 recurrences were observed. In addition, 99 patients progressed to MIBC or mBC. The median follow-up time for the complete cohort was 59 months (interquartile range (IQR): 38-67 months) and varied per risk group due to the extended inclusion of T1 patients in the UroLife study. Median follow-up times for low, intermediate, high, and very high risk NMIBC were respectively 62, 62, 46, and 31 months. The median age at initial diagnosis was 67 years (range: 18-80 years); 79% of the population was male.

**Table 1.**
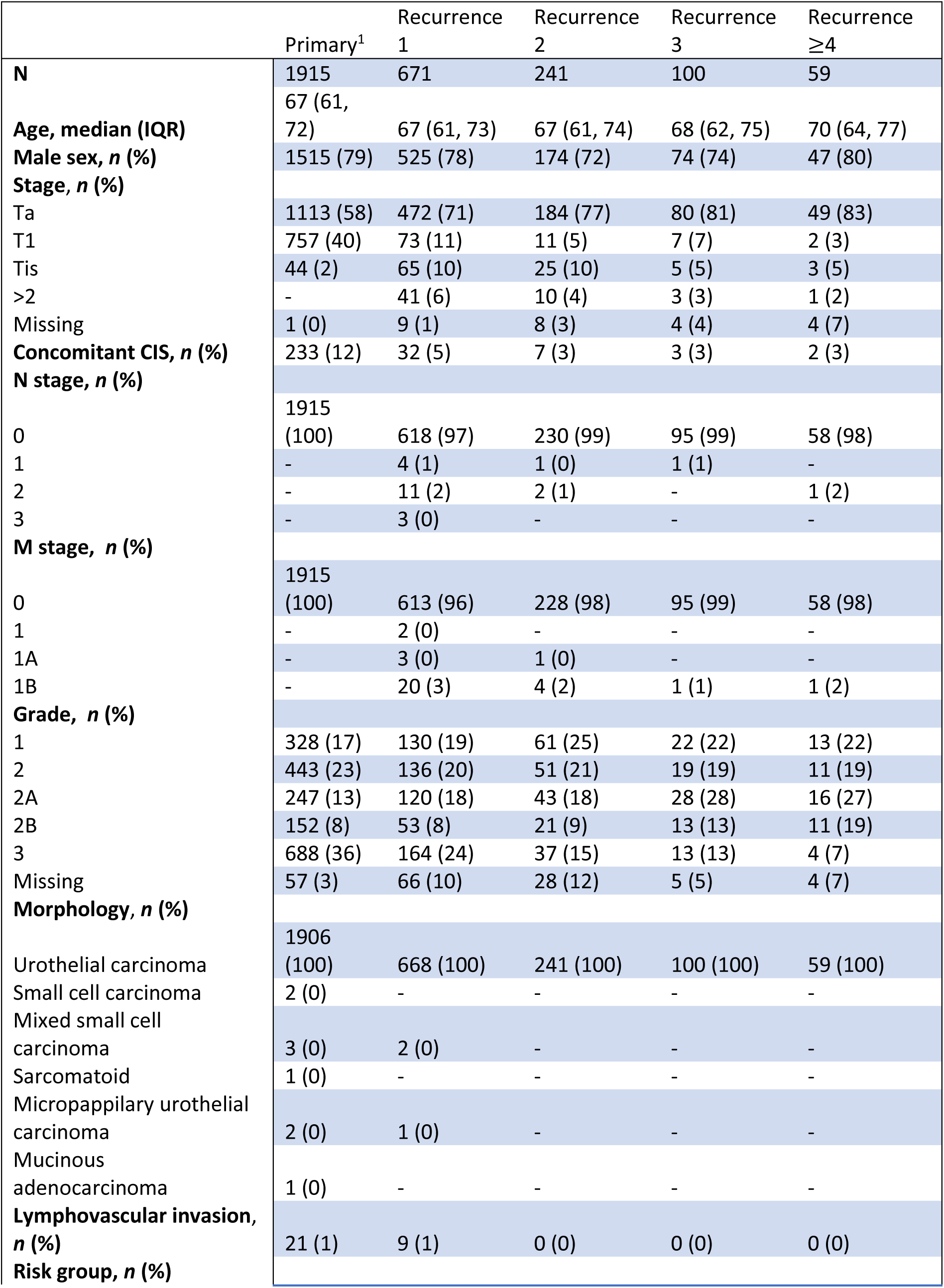
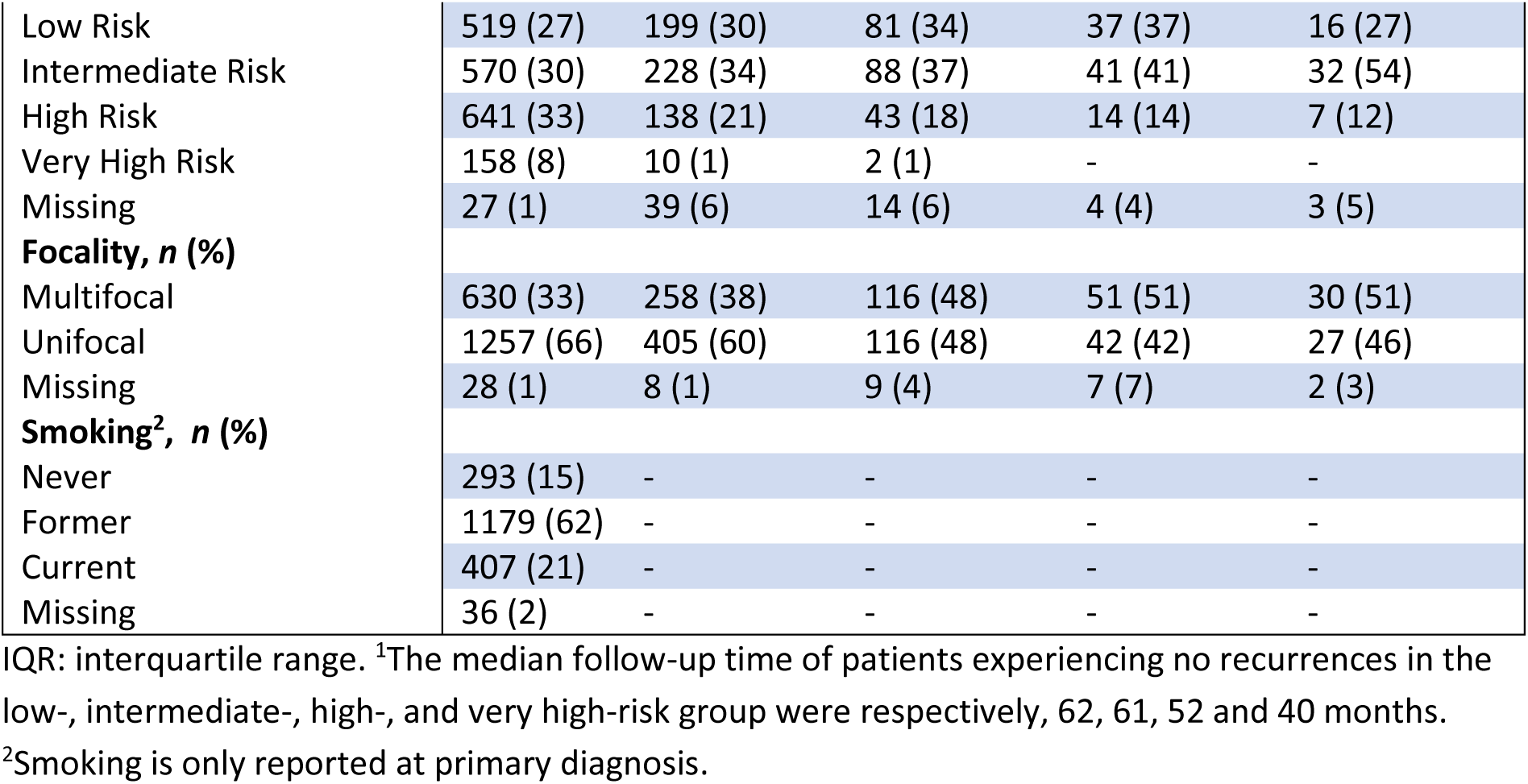
Characteristics of all observed tumours in UroLife and the Nijmegen Bladder Cancer Study. Tumour characteristics are presented for all primary tumours (i.e., the complete cohort at time of diagnosis), and all first, second, third and later recurrences, thereafter. Due to missing values, clinical characteristics were not available for all tumours.

Details of the treatment of primary tumours, categorized by risk group, are provided in **Table 2**. Single postoperative instillation of chemotherapy was applied in 61% of low risk patients. High- and very high risk papillary tumours underwent a re-TURBT in 72% and 82% of cases. Induction BCG therapy was administered to 74% of high risk and 68% of very high risk primary tumours. In total, 44 patients underwent radical cystectomy for the primary tumour, and another 85 patients underwent radical cystectomy later during follow-up: respectively 66 (10%) and 43 (27%) patients in the high- and very high risk group were at some point treated with radical cystectomy.

**Table 2.** Treatment of primary tumours, stratified by risk group.

|  | Low risk | Intermediate risk | High risk | Very high risk |
| --- | --- | --- | --- | --- |
| **N** | 519 | 570 | 641 | 158 |
| **re-TURBT\*, n (%)** | 38 (7) | 121 (21) | 455 (71) | 122 (77) |
| **Radical cystectomy, n (%)** | 3 (1) | 10 (2) | 22 (3) | 9 (6) |
| **Single post-operative instillation of chemotherapy, n (%)** | 317 (61) | 345 (61) | 225 (35) | 42 (27) |
| *Mitomycine, n (%)*<sup>x</sup> | 205 (65) | 258 (75) | 164 (73) | 32 (76) |
| *Epirubicine, n (%)* | 103 (32) | 59 (17) | 52 (23) | 7 (17) |
| *Gemcitabine, n (%)* | - | - | - | - |
| *Unspecified, n (%)* | 9 (3) | 28 (8) | 9 (4) | 3 (7) |
| **Adjuvant chemotherapy, n (%)** | 107 (21) | 185 (32) | 139 (22) | 18 (11) |
| *Mitomycine, n (%)*<sup>x</sup> | 71 (66) | 153 (83) | 82 (60) | 8 (44) |
| *No. instillations, median (IQR)* | 9 (7, 9) | 9 (7, 9) | 8 (5, 9) | 8 (4, 13) |
| *Epirubicine, n (%)* | 33 (31) | 17 (9) | 14 (10) | 1 (6) |
| *No. instillations, median (IQR)* | 2 (1, 7) | 6 (2, 8) | 1 (1, 9) | 1 (1, 1) |
| *Gemcitabine, n (%)* | - | 6 (3) | 39 (28) | 8 (44) |
| *No. instillations, median (IQR)* | - | 6 (6, 8) | 6 (5, 11) | 7 (6, 9) |
| *Unspecified, n (%)* | 3 (3) | 9 (5) | 2 (1) | 1 (6) |
| *No. instillations, median (IQR)* | 11 (6, 12) | 9 (7, 9) | - | - |
| **Induction BCG<sup>+</sup>, n (%)** | 3 (1) | 85 (15) | 472 (74) | 108 (68) |
| *No. instillations, median (IQR)* | 6 (6, 6) | 6 (6, 6) | 6 (6, 6) | 6 (6, 6) |
| **Maintenance BCG<sup>+</sup>, n (%)** | 3 (1) | 70 (12) | 317 (49) | 80 (51) |
| *No. instillations, median (IQR)* | 10 (8, 12) | 10 (9, 15) | 9 (6, 15) | 9 (3, 12) |

\*re-TURBT was performed for 446 (72%) out of 618 papillary high risk tumours and 110 (82%) out of 134 papillary very high risk tumours. <sup>x</sup>Percentage of patients receiving a type of single instillation- or adjuvant chemotherapy is reported relative to patient subgroup. <sup>+</sup>Induction BCG and maintenance BCG are defined in Supplementary Table 4.

### Incidence and timing of recurrences

Of all observed recurrences, 671 (63%) were first recurrences. Among patients experiencing at least one recurrence, 241 had at least one additional recurrence (i.e., two recurrences), 100 had at least three recurrences and 39 had four recurrences or more.

KM curves for first to fourth recurrence are displayed in **Figure 1** (KM risks and 95% confidence intervals in **Supplementary Table 2)**. The 1-year, 3-year, and 5-year KM risks of a first recurrence in the total population were 15%, 31%, and 39%, respectively. We observed an increased risk for subsequent recurrences: the 1-year, 3-year, and 5-year conditional KM recurrence risks of a second recurrence after the first recurrence were 24%, 47%, and 52%, whereas the 1-year, 3-year, and 5-year conditional KM recurrence risks of a third recurrence were even higher: 28%, 55%, and 60%, respectively.

**Figure 1.**
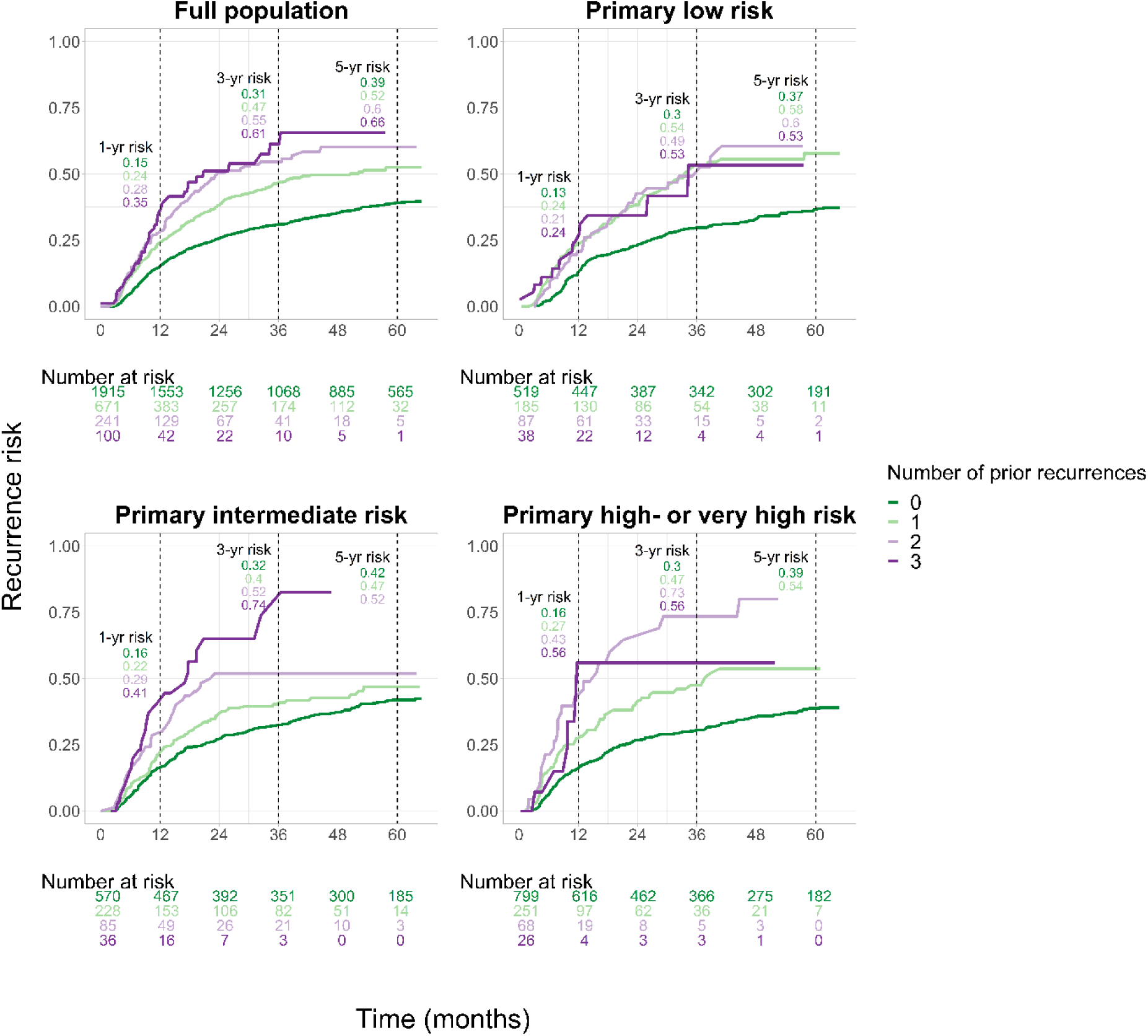
Recurrence risk of NMIBC for the first to fourth recurrence, for complete cohort and stratified by risk group of primary tumour. Recurrence risk is reported as Kaplan-Meier (KM) risks, based on time to recurrence since tumour-free status. For all KM curves, the 1-, 3-, and 5-year risks are included in the figure, and are reported with 95% confidence intervals in Supplementary Table 2. Note that some numbers at risk of risk groups may not add up to numbers of the full population due to missing values in risk groups.

The 3-year KM risk of a first recurrence was similar for patients initially diagnosed with low-, intermediate- and high-risk NMIBC: 30%, 32% and 30%, respectively. The 3-year KM risks of a second recurrence after the first in the low-, intermediate and high-risk group were respectively 54%, 40% and 47%.

We found similar recurrence risks of first to fourth recurrence after stratifying by sex and year of diagnosis of primary tumour (**Supplementary Figure 1**). The impact of (indication of) treatment on recurrence patterns is visualized in **Supplementary Figure 2**. Patients with primary low risk NMIBC who were treated with a single postoperative chemotherapy instillation had comparable rates of subsequent recurrences compared to those without. Primary high risk patients who received a re-TURBT had lower rates of a first recurrence compared to patients without re-TURBT, and high risk patients who received BCG induction had lower rates of first recurrence compared to patients not receiving BCG induction therapy.

### Patterns of recurrences

Figure 2 displays all transitions between risk groups (low risk, intermediate risk, high or very high risk, MIBC/mBC) between the primary tumour and subsequent recurrences. A recurrence after a low, intermediate or high/very high risk tumour (either primary or recurrence) was again of low-, intermediate- or high-risk in 74%, 62% and 44% of cases. Concordance was even higher when focusing on grade and stage: a recurrence after low grade NMIBC (grade 1, 2, or 2A) was also of low grade in 91% of cases, while a recurrence after high grade NMIBC was also of high grade in 80% of cases. As for stage, 87% of recurrences after a Ta tumour were again of stage Ta. By contrast, only 25% of recurrences after a T1 tumour were again stage T1, while respectively 38%, 15% and 19% of recurrences were Ta, (concomitant) CIS, or MIBC/mBC. Finally, 52% of recurrences after (concomitant) CIS were again (concomitant) CIS (**Supplementary Figures 3 and 4**).Of patients with primary low-risk NMIBC, 2.9%, 7.7%, and 10.4% developed a recurrence of intermediate or higher risk within one year, three years, and five years after diagnosis, respectively.

**Figure 2.**
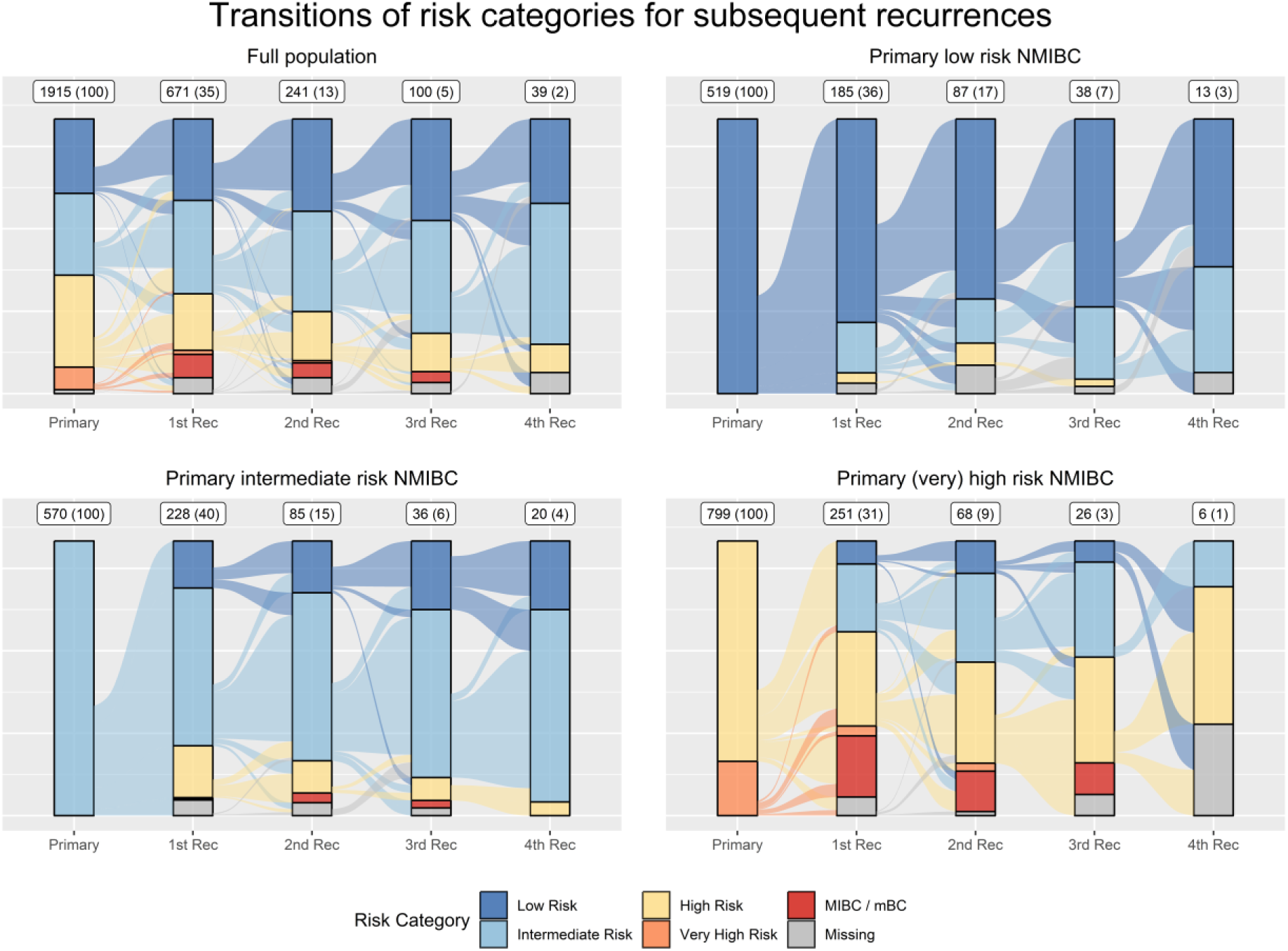
Alluvial diagram describing transitions of risk groups and progression within patients across subsequent recurrences, for complete cohort and stratified by risk group of primary tumour. The vertical bars indicate the distribution of risk groups of tumours for the primary tumour and first to fourth recurrences. The numbers on top of each bar indicate the numbers of tumours and proportion in this (sub-)population. The stream fields between the blocks indicate the individual changes in risk group for subsequent recurrences. We note that risk group was not available for all tumours due to missing values.

Data on tumour location was available for 1,492 of the 1,915 primary tumours and 667 of the 1,071 recurrences. Alluvial plots that visualise the transitions of tumour locations are presented in **Supplementary Figure 5**. In 78% of recurrences with tumour location available, the initial and recurrent tumour were of the same coded site. For the subgroup of patients who received a re-TURBT of an initial high- or very high risk tumour, 95 out of 140 recurrences (68%) with known tumour location were of the same location as the previous tumour, and in the subgroup of patients without re-TURBT, 52 out of 72 recurrences (72%) were of the same location.

In total, 99 patients progressed to MIBC or mBC. The disease course of these patients before progression is visualised in Figure 3. Among the patients who progressed to MIBC or mBC, one patient was diagnosed with primary low risk NMIBC, 9 with primary intermediate risk NMIBC, and 89 with primary high or very high risk NMIBC. The patient with primary low risk NMIBC had a concomitant Ta + CIS recurrence before progressing to a T2 tumour. Among the 9 patients with primary intermediate risk NMIBC, eight patients had a recurrence before progression. Six of them had a high risk grade 2B or 3 recurrence before progressing to MIBC or mBC; the other two experienced an intermediate risk recurrence before progression. Among the 10 patients with primary low- or intermediate risk NMIBC who progressed to MIBC or mBC, four patients underwent a re-TURT after TURBT of primary NMIBC. Seven patients out of 10 were at some point treated with bacillus Calmette-Guérin (BCG), and eight out of 10 were treated with intravesical chemotherapy.

**Figure 3.**
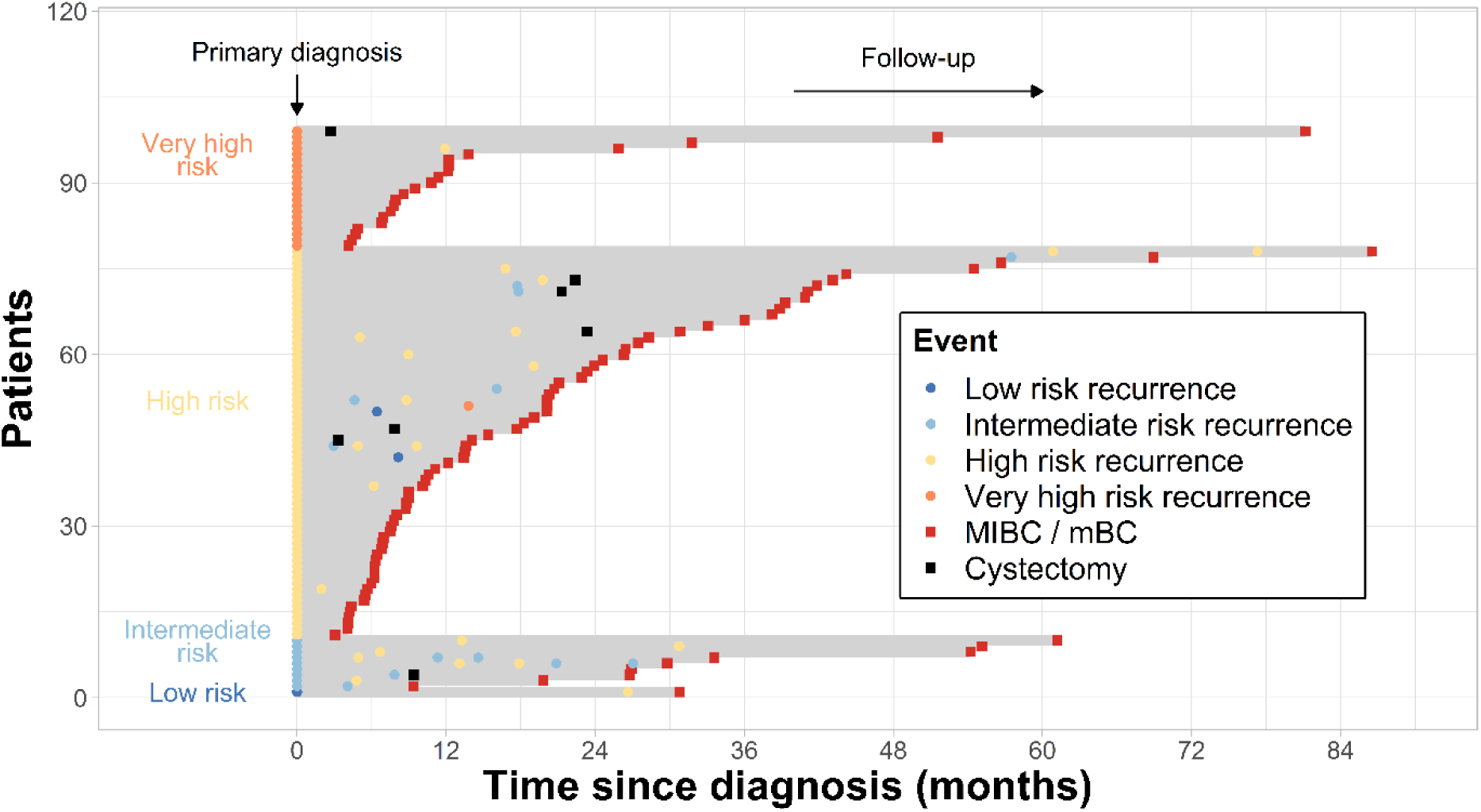
Swimmer plot describing the timing of recurrences, cystectomy and progression to MIBC or mBC for patients who progressed to MIBC or mBC. Patients are categorized based on EAU group of primary tumour and ordered on the y-axis based on duration of follow-up time (depicted as grey line). All observed recurrences or cystectomy before progression to MIBC or mBC are plotted on the relevant y-value of the individual.

Among the 89 patients with primary high or very high risk NMIBC, 65 (73%) experienced progression to stage 2 or higher, 29 (33%) were positive for lymph node metastasis and 35 (39%) experienced distant metastasis. Only 19 (21%) patients experienced a recurrence before progression, of which 13 (15%) were high- or very high-risk. However, it should be noted that 9 of the 89 patients were never tumour-free after primary (concomitant) CIS, and therefore never at risk of recurrence. Of these 89 patients, 42 underwent a re-TURBT after the TURBT of the primary tumour; 73 patients were at some point treated with BCG, 37 were treated with multiple intravesical chemotherapy instillations for the primary tumour, and 4 patients were never treated with intravesical chemo- or immunotherapy.

## Discussion

In this large contemporary NMIBC cohort, we investigated patterns of multiple recurrences and progression. We extensively described the transitions, timings and characteristics of subsequent recurrences, as well as the course of the disease before progression to MIBC or mBC. Our findings provide novel insights into the course of multiple recurrences in NMIBC under the current guidelines, which can be used for future studies into surveillance schedules and facilitate research into NMIBC recurrence mechanisms.

One pivotal outcome of our research is the observation that recurring tumours often manifest similar clinicopathological characteristics, such as similar grade, stage, and risk group. This phenomenon was previously reported in literature (15, 22), but now quantitatively substantiated by our study. Notably, among the patients with primary low- or intermediate-risk NMIBC progressing to MIBC or mBC, we observed a pattern in which progression was preceded by a high- or very high risk recurrence in 70% of the patients. Conversely, primary high- or very high-risk NMIBC patients often exhibited a direct progression to MIBC or mBC, with only 21% reporting any recurrence before progression. We also observed that recurrence risks increased with the number of previous recurrences, in line with current NMIBC risk calculators (5, 6).

Most diagnoses of primary and recurrent bladder cancer are based on invasive and costly cystoscopy, prompting the question whether patients could be monitored using a more relaxed schedule or through urine markers instead. Based on our data, we cannot evaluate possible effects of changes in surveillance schedules on NMIBC prognosis, as it remains unknown how tumours develop if detected at a later point in time. We found that high grade recurrences often precede progression to MIBC or mBC after low- or intermediate risk NMIBC. Urine markers work best in detecting high grade recurrences (23), suggesting that for this subgroup, replacing cystoscopy checks with urine markers could control MIBC/mBC risks by timely detection of high risk recurrences. Nonetheless, comprehensive population-based data and increased understanding of the biological process of progression is needed to assess the prognostic implications of substituting cystoscopy with urine markers.

Prior studies have explored patterns of subsequent NMIBC recurrences, focusing on recurrence mechanisms (15), statistical methods (14) or risk factors for multiple recurrences (13, 16–18). No study reported the association between NMIBC risk groups and patterns of subsequent NMIBC or MIBC/mBC development. Despite having a relatively long median follow-up time in the combined dataset of UroLife and NBCS, other studies generally report higher recurrence rates (see **Supplementary Table 4**). This may be due to our stricter definition of NMIBC recurrences, which requires a tumour-free status before a recurrence, but also due to differences in patient populations or intensification of NMIBC treatment over time.

Our study’s primary strength lies in the size and quality of the combined datasets from UroLife and NBCS. Clinical data were carefully collected in consultation with urologists and experts in bladder cancer, enabling the in-depth exploration of recurrence patterns. The size of the dataset enabled the stratification on various clinical characteristics of patients and visual presentation of tumour characteristics up to the fourth recurrence. However, we recognise some limitations of the study. The inclusion in the last four years of the UroLife study was restricted to T1 patients, resulting in a relatively short follow-up time for this subgroup. Our population-based cohort was not treated in full concordance with the current EAU guidelines. For example, only 61% of low- or intermediate risk patients received a single post-operative instillation of chemotherapy. However, this is in line with earlier reports on adherence to guidelines(12, 24, 25). Thus, our study offers a realistic insight into patterns of multiple recurrences in a contemporary NMIBC population and differences in these patterns according to treatment. We were unable to incorporate tumour size in our definition of NMIBC risk groups. As a result, a small percentage of tumours may be assigned to a lower risk group compared to assignment based on the EAU risk group classification. Also, prior recurrence was omitted as risk factor, to allow for a direct comparison of intrinsic pathological characteristics among subsequent tumours. Finally, this study was limited to two Dutch bladder cancer studies, and replication in other populations is needed to externally validate our findings.

## Conclusions

Using a large contemporary cohort, we revealed key patterns in NMIBC recurrences. We demonstrated that recurring NMIBC tumours often manifest similar clinicopathological characteristics. Over half of patients with primary low- or intermediate risk NMIBC who progress to MIBC or mBC experience a high- or very high risk recurrence before progression, whereas primary high risk NMIBC patients often exhibit direct progression to MIBC/mBC. The results of this study can assist in the optimization of the design of future studies into factors that affect NMIBC outcomes. Also, our study highlights relevant areas for further research, including further research into specific patient subgroups with distinct recurrence patterns.

## Data Availability

All data produced in the present study are available upon reasonable request to the authors

## Acknowledgement

The authors thank all the patients who participated in UroLife and thank the following hospitals for their involvement in recruitment for the UroLife study: Amphia Ziekenhuis, Breda/Oosterhout (D.K.E. van der Schoot); Ziekenhuis Bernhoven, Uden (A.Q.H.J. Niemer); Canisius-Wilhelmina Ziekenhuis, Nijmegen (D.M. Somford); Catharina Ziekenhuis, Eindhoven (E.L. Koldewijn); Deventer Ziekenhuis, Deventer (P.L.M. van den Tillaar); Elkerliek Ziekenhuis, Helmond (E.W. Stapper †, P.J. van Hest); Gelre Ziekenhuizen, Apeldoorn/Zutphen (D.M. Bochove-Overgaauw); Isala Klinieken, Zwolle (E. te Slaa); Jeroen Bosch Ziekenhuis, ‘s-Hertogenbosch (S. van der Meer); Meander Medisch Centrum, Amersfoort (F.S. van Rey); Medisch Spectrum Twente, Enschede (M. Asselman); Maxima Medisch Centrum, Veldhoven/Eindhoven (L.M.C.L. Fossion, K. de Laet); Maasziekenhuis Pantein, Boxmeer (E. van Boven); Radboudumc, Nijmegen; Rijnstate, Arnhem/Velp/Zevenaar (C.J. Wijburg); Slingeland Ziekenhuis, Doetinchem (A.D.H. Geboers); St. Anna Ziekenhuis, Geldrop (A. Sonneveld); Elisabeth-TweeSteden Ziekenhuis, Tilburg/Waalwijk (P.J.M. Kil, B.P. Wijsman); St. Jansdal Ziekenhuis, Harderwijk (W.J. Kniestedt); VieCuri, Venlo (G. Yurdakul, A.H.P. Meier); Ziekenhuis Gelderse Vallei, Ede (M.D.H. Kortleve); Ziekenhuisgroep Twente, Almelo/Hengelo (E.B. Cornel). The authors thank the registration team of the Netherlands Comprehensive Cancer Organisation (IKNL) for the collection of data for the Netherlands Cancer Registry as well as IKNL staff for scientific advice.

