## Supplemental Information for "Recurrence patterns in a large contemporary cohort of patients with non-muscle invasive bladder cancer"

*[*]These authors share senior authorship*

### Supplementary Figures

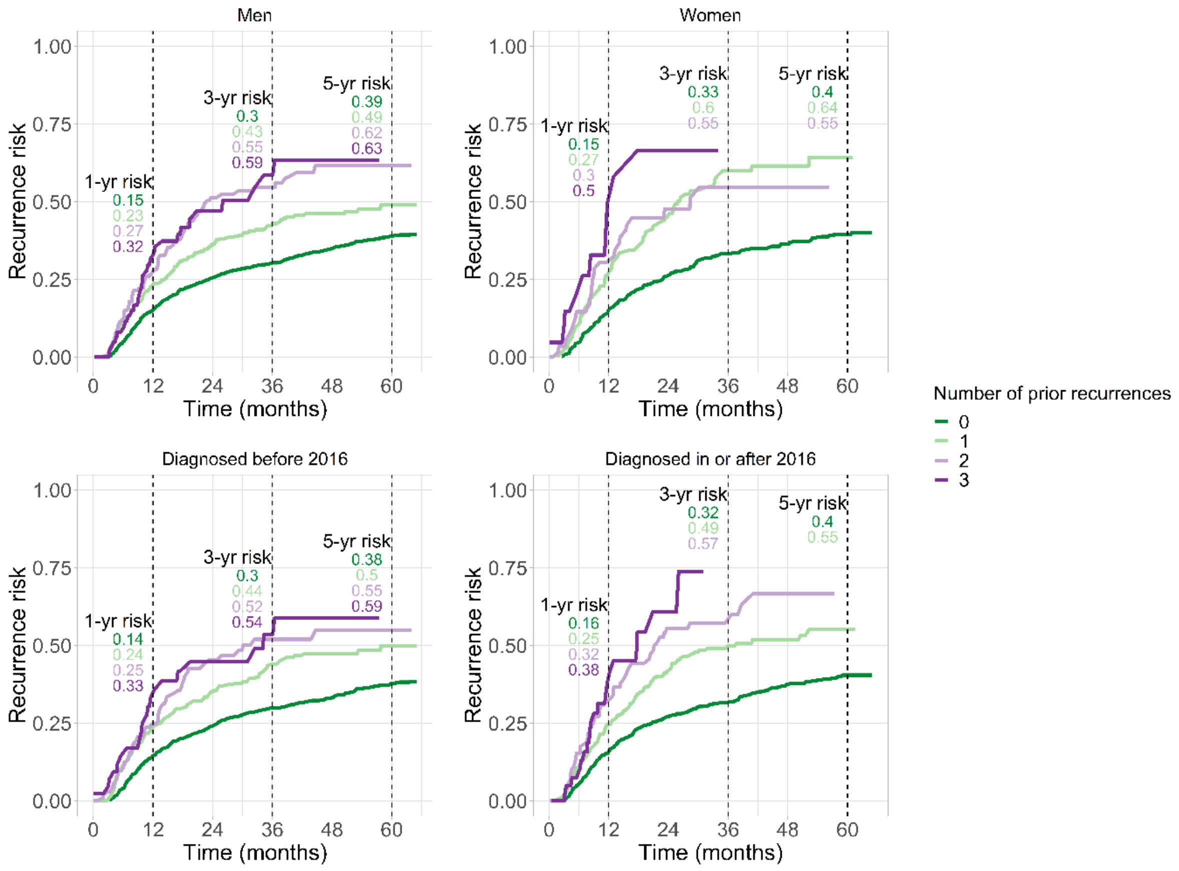


**Supplementary Figure 1. Recurrence risk for the first to fourth recurrence, stratified by sex and time of diagnosis.** Recurrence risk was evaluated based on time since achieving a tumour-free status after the previous NMIBC tumour.

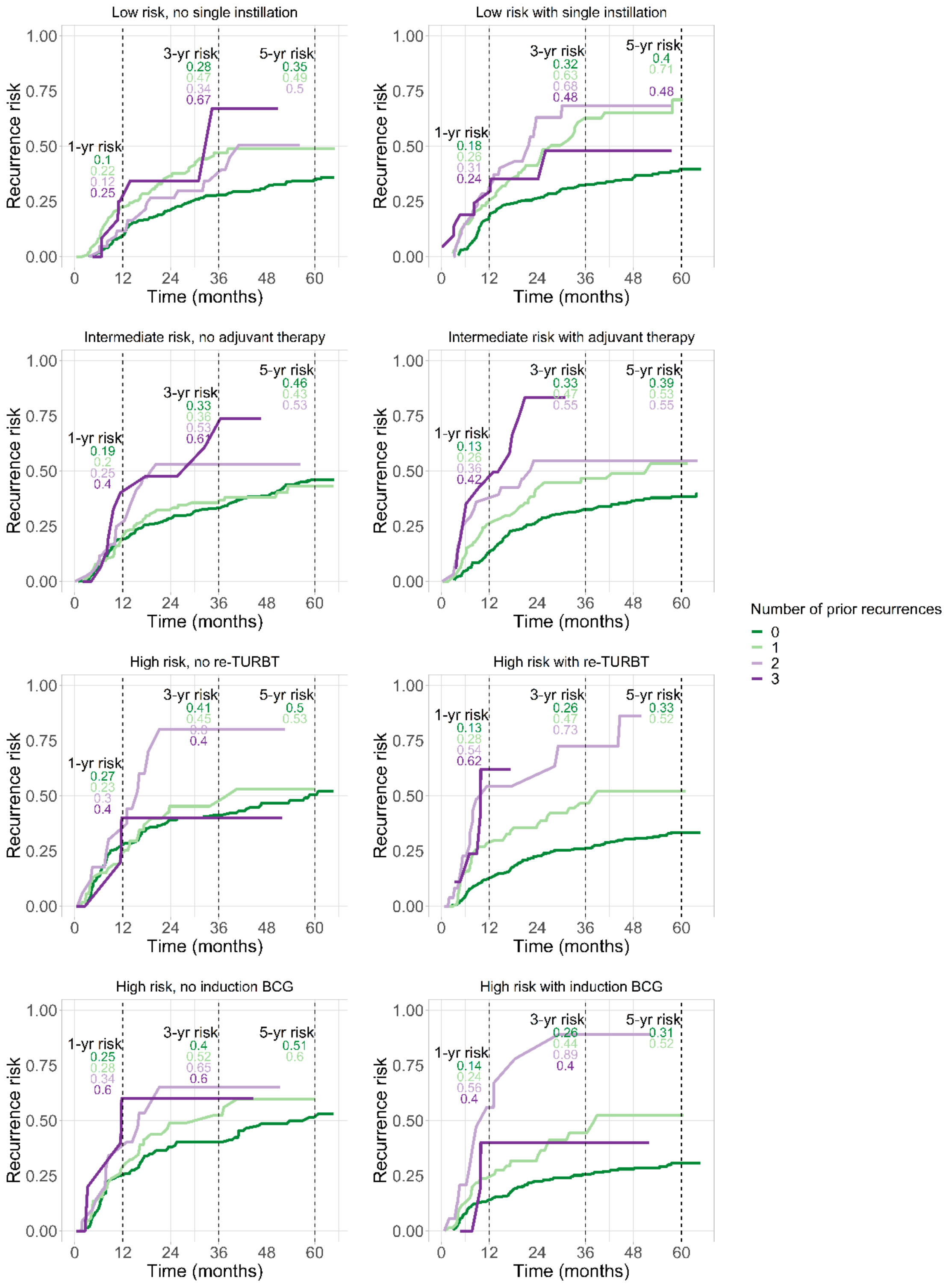


**Supplementary Figure 2. Recurrence risk for the first to fourth recurrence, stratified by risk category and relevant treatment.** The top row compares first to fourth recurrence risk in primary low risk NMIBC, stratified by treatment with single immediate postoperative chemotherapy instillation. The second row compares recurrence risks for primary intermediate risk NMIBC patients, stratified by adjuvant (either chemo- or BCG-)therapy. The third and fourth row compare recurrence rates in the high risk group, stratified by treatment with re-TURBT and induction BCG for primary tumour, respectively. Recurrence risk was evaluated based on time since achieving a tumour-free status after the previous NMIBC tumour.

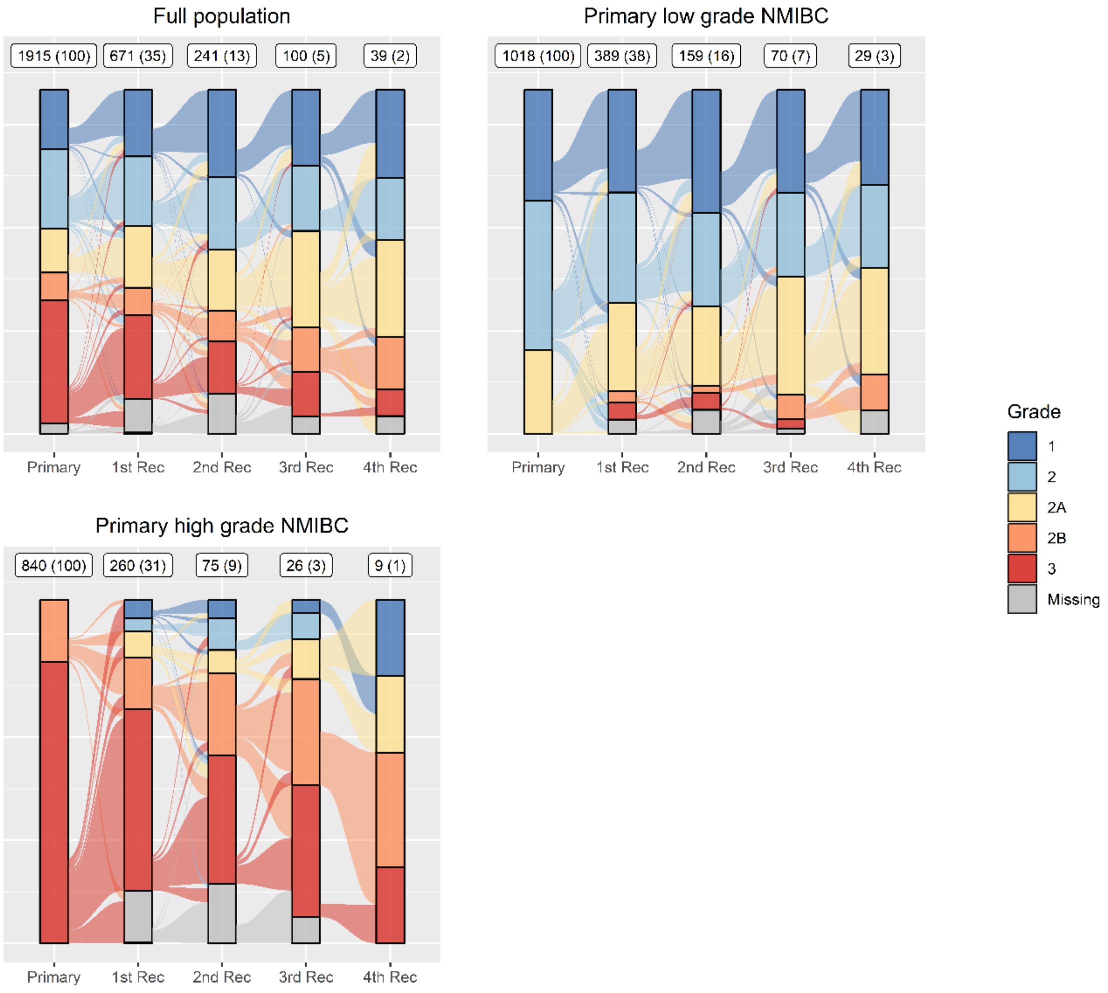


**Supplementary Figure 3. Transitions of grade within patients across subsequent recurrences, for full population and stratified by grade of primary tumour.** The vertical bars indicate the distribution of grade of tumours for the primary tumour and first to fourth recurrences. The numbers on top of each bar indicate the numbers of tumours and proportion for the primary tumour or recurrence in this (sub-)population. Grade was not available for all tumours due to missing values.

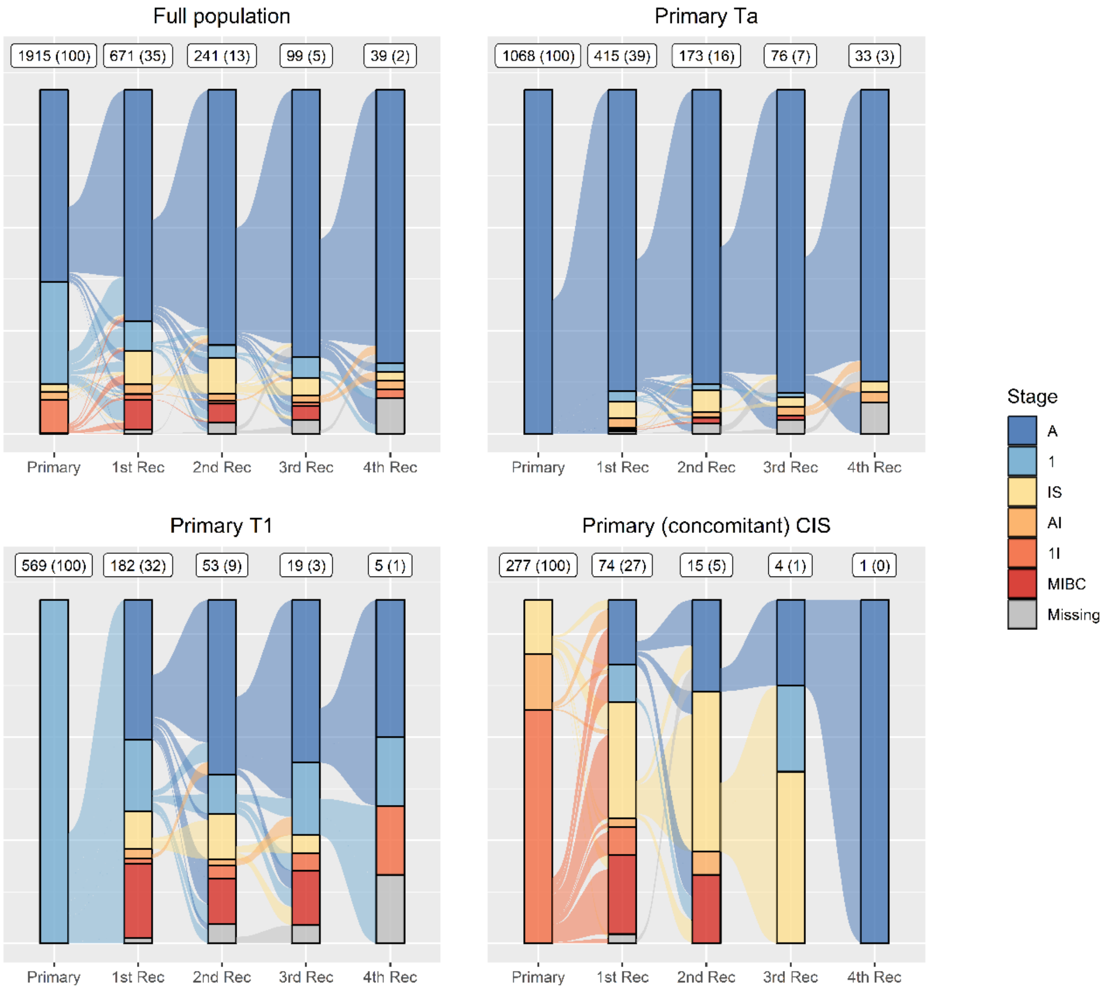


**Supplementary Figure 4. Transitions of stage within patients across subsequent recurrences, for full population and stratified on stage of primary tumour.** IS: carcinoma in situ. AI: concomitant Ta + Tis. 1I: concomitant T1 + Tis. The vertical bars indicate the distribution of stage of tumours for the primary tumour and first to fourth recurrences. The numbers on top of each bar indicate the numbers of tumours and proportion for the primary tumour or recurrence in this (sub-)population. Stage was not available for all tumours due to missing values.

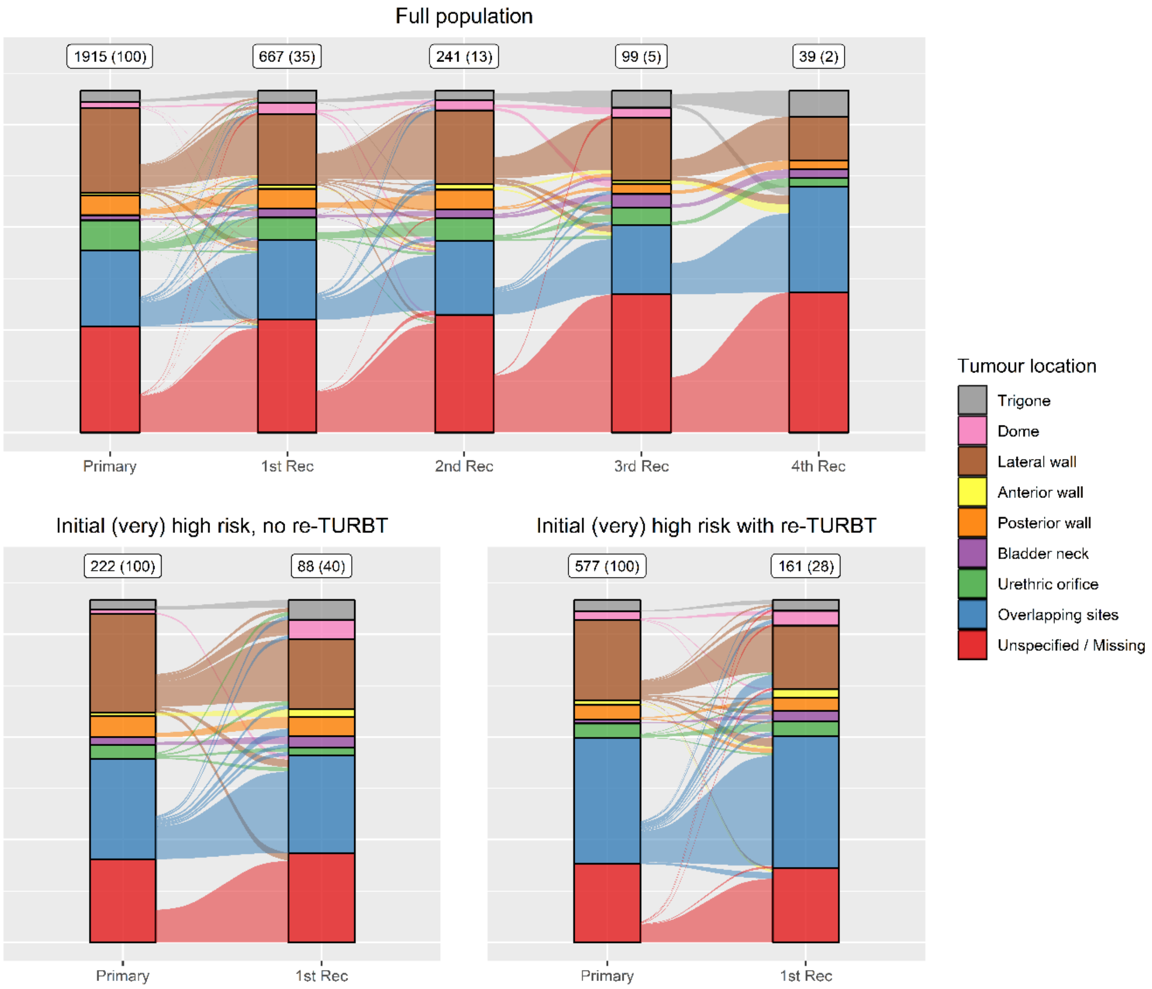


**Supplementary Figure 5. Transitions of tumour location within patients across subsequent recurrences.** The vertical bars indicate the distribution of tumour location for the primary tumour and first (to fourth) recurrences. The numbers on top of each bar indicate the numbers of tumours and proportion for the primary tumour or recurrence in this (sub-)population. The top row presents the transitions in tumour location for the full population; the bottom row presents the transitions in tumour location for primary (very) high risk patients, stratified by re-TURBT for primary tumour. Note that tumour location was not available for all tumours due to unspecified and missing values.

### Supplementary Tables

**Supplementary Table 1. Definitions of risk groups used in main article, obtained from EAU prognostic risk categories (1).**

| Risk Group | |
|---|---|
| **Low risk** | • Single, TaT1 LG/G1 tumours without CIS in patients ≤70 years<br>• Ta LG/G1 tumours without CIS with at most ONE of the additional risk factors* |
| **Intermediate risk** | Tumours without CIS that are not included in low, high or very high-risk groups |
| **High risk** | • All T1 HG/G3 tumours without CIS, EXCEPT those included in the very high-risk group<br>• All CIS tumours, EXCEPT those included in the very high-risk group<br>• Ta HG/G3 no CIS with 2 risk factors<br>• T1 G2 no CIS with at least 1 risk factor |
| **Very high risk⁺** | • T1G2 with CIS and 2 risk factors<br>• T1 HG/G3 with CIS and at least 1 risk factor |

LG denotes grade G1, G2 or G2a, HG denotes grade G2b or G3. *Additional risk factors: Age > 70; Multifocal tumour. ⁺Presence of histological subtypes or lymphovascular invasion (only available for UroLife study) were coded as very high risk.

**Supplementary Table 2. Kaplan-Meier risk of first to fourth recurrence, for the total population and stratified by risk group of primary tumour, including 95% confidence intervals.** Recurrence risk was evaluated based on time since achieving a tumour-free status after the previous NMIBC tumour.

| | Rec | 1 year | 2 years | 3 years | 4 years | 5 years |
|---|---|---|---|---|---|---|
| Total population | 1 | 0.15 (0.14, 0.17) | 0.26 (0.24, 0.28) | 0.31 (0.29, 0.33) | 0.35 (0.33, 0.38) | 0.39 (0.37, 0.41) |
| | 2 | 0.24 (0.21, 0.28) | 0.38 (0.34, 0.42) | 0.47 (0.42, 0.51) | 0.5 (0.45, 0.54) | 0.52 (0.47, 0.57) |
| | 3 | 0.28 (0.22, 0.34) | 0.5 (0.42, 0.57) | 0.55 (0.46, 0.62) | 0.6 (0.51, 0.68) | 0.6 (0.51, 0.68) |
| | 4 | 0.35 (0.23, 0.46) | 0.51 (0.37, 0.62) | 0.61 (0.44, 0.73) | 0.66 (0.47, 0.78) | 0.66 (0.47, 0.78) |
| Low risk | 1 | 0.13 (0.1, 0.16) | 0.23 (0.19, 0.27) | 0.3 (0.26, 0.34) | 0.33 (0.29, 0.37) | 0.37 (0.32, 0.41) |
| | 2 | 0.24 (0.17, 0.3) | 0.38 (0.31, 0.45) | 0.54 (0.45, 0.61) | 0.56 (0.46, 0.63) | 0.58 (0.48, 0.66) |
| | 3 | 0.21 (0.11, 0.29) | 0.43 (0.3, 0.53) | 0.49 (0.35, 0.6) | 0.6 (0.42, 0.73) | 0.6 (0.42, 0.73) |
| | 4 | 0.24 (0.08, 0.37) | 0.34 (0.15, 0.49) | 0.53 (0.19, 0.73) | 0.53 (0.19, 0.73) | 0.53 (0.19, 0.73) |
| Intermediate risk | 1 | 0.16 (0.13, 0.19) | 0.27 (0.23, 0.31) | 0.32 (0.28, 0.36) | 0.37 (0.33, 0.41) | 0.42 (0.38, 0.46) |
| | 2 | 0.22 (0.16, 0.28) | 0.36 (0.29, 0.43) | 0.4 (0.33, 0.47) | 0.43 (0.35, 0.49) | 0.47 (0.38, 0.54) |
| | 3 | 0.29 (0.17, 0.38) | 0.52 (0.38, 0.62) | 0.52 (0.38, 0.62) | 0.52 (0.38, 0.62) | 0.52 (0.38, 0.62) |
| | 4 | 0.41 (0.2, 0.56) | 0.65 (0.41, 0.79) | 0.74 (0.43, 0.88) | - | - |
| High- or very high risk | 1 | 0.16 (0.13, 0.19) | 0.27 (0.23, 0.3) | 0.3 (0.27, 0.34) | 0.36 (0.32, 0.39) | 0.39 (0.35, 0.43) |
| | 2 | 0.27 (0.2, 0.34) | 0.41 (0.32, 0.49) | 0.47 (0.38, 0.56) | 0.54 (0.43, 0.62) | 0.54 (0.43, 0.62) |
| | 3 | 0.43 (0.25, 0.56) | 0.65 (0.42, 0.78) | 0.73 (0.5, 0.86) | 0.8 (0.54, 0.91) | - |
| | 4 | 0.56 (0.11, 0.78) | 0.56 (0.11, 0.78) | 0.56 (0.11, 0.78) | 0.56 (0.11, 0.78) | - |

**Supplementary Table 3. Baseline characteristics and treatment of primary tumours for the 99 patients that progressed to MIBC / mBC.**

| | Low risk | Intermediate risk | High risk | Very high risk |
|---|---|---|---|---|
| ***Baseline characteristics*** | | | | |
| **N** | 1 | 9 | 68 | 21 |
| **Age, median (IQR)** | 64 (64, 64) | 68 (64, 73) | 68 (62, 73) | 71 (66, 75) |
| **Male sex** | 1 (100) | 9 (100) | 52 (76) | 17 (81) |
| **Stage** | | | | |
| Ta | 1 (100) | 8 (89) | - | - |
| T1 | - | 1 (11) | 51 (75) | 8 (38) |
| Ta + Tis | - | - | 2 (3) | - |
| T1 + Tis | - | - | 10 (15) | 13 (62) |
| Tis | - | - | 5 (7) | - |
| **Grade** | | | | |
| 1 | - | 1 (11) | - | - |
| 2 | - | 4 (44) | 7 (10) | 1 (5) |
| 2A | 1 (100) | - | - | - |
| 2B | - | 1 (11) | 4 (6) | - |
| 3 | - | 3 (33) | 51 (75) | 17 (81) |
| Missing | - | - | 6 (9) | 3 (14) |
| ***Treatment*** | | | | |
| **re-TURBT** | 0 (0) | 4 (44) | 34 (50) | 8 (38) |
| **Radical Cystectomy*** | 0 (0) | 1 (11) | 9 (13) | 2 (10) |
| **Peri-operative Chemo** | 0 (0) | 5 (56) | 16 (24) | 6 (29) |
| Mitomycine+ | - | 3 (60) | 14 (88) | 4 (67) |
| Epirubicine+ | - | 1 (20) | 1 (6) | 1 (17) |
| Gemcitabine+ | - | - | - | - |
| Unspecified+ | - | 1 (20) | 1 (6) | 1 (17) |
| **Adjuvant Chemo** | 0 (0) | 4 (44) | 15 (22) | 3 (14) |
| Mitomycine+ | - | 3 (75) | 8 (57) | 1 (33) |
| Nr instillations, median (IQR) | - | 9 (5, 9) | 8 (7, 9) | 5 (5, 5) |
| Epirubicine+ | - | 1 (25) | 2 (14) | - |
| Nr instillations, median (IQR) | - | - | 1 (1, 1) | - |
| Gemcitabine+ | - | - | 4 (29) | 2 (67) |
| Nr instillations, median (IQR) | - | - | 6 (4, 10) | 9 (9, 9) |
| **Induction BCG** | 0 (0) | 2 (22) | 47 (69) | 9 (43) |
| **Nr instillations, median (IQR)** | - | 6 (6, 6) | 6 (6, 6) | 6 (6, 6) |
| **Maintenance BCG** | 0 (0) | 1 (11) | 21 (31) | 5 (24) |
| **Nr instillations, median (IQR)** | - | - | 9 (6, 11) | 3 (2, 4) |

*Note that radical cystectomy only counted if it occurred before progression to MIBC / mBC. +The percentage of individuals receiving the type of instillation are reported relative to patient subgroup.

**Supplementary Table 4. Overview of data used in studies into multiple NMIBC recurrences**.

| Reference | Data collection | Sample size | Median follow-up time (months) | Number of first recurrences (%) | Total number of recurrences (avg. per person) |
|---|---|---|---|---|---|
| **Current study: UroLife + NBCS** | 2011-2021 | 1,915 | 59 | 671 (35) | 1,071 (0.55) |
| **Bryan et al. (2)** | 1999-2004 | 169 | 34 | 79 (46) | 198 (1.17) |
| **Smedinga et al. (3)** | 1990-2012 | 531 | 53* | 237 (45) | 656 (1.24) |
| **Waseda et al. (4)** | 2000-2016 | 585 | 41 | 253 (43) | 475 (0.8) |
| **Sharma et al. (5)** | 2015-2019 | 291 | 38 | 137 (47) | ≥234 (0.8)[x] |
| **Simon et al. (6)** | 1986-2010 | 470 | 86 | 251 (53) | 759 (1.61) |

*Follow-up was only directly reported for patients experiencing no recurrence. [x]At least 234 recurrences were recorded in this study (total number not available).

**Supplementary Table 5. Definitions of induction BCG and maintenance BCG in the UroLife study and Nijmegen Bladder Cancer Study.**

| | Definition |
|---|---|
| **Induction BCG** | At least one of the following holds:<br>• Tumour is treated with at least 5 BCG instillations<br>• Tumour is treated with an unknown number of instillations, but more than 26 days between first and last BCG instillation |
| **Maintenance BCG** | Both the following hold:<br>• Received induction BCG<br>• Received at least 7 instillations OR received an unknown number of instillations, with at least 70 days between first and last BCG instillation |
